# Study protocol for a prospective multicenter longitudinal cohort study on progression of moderate aortic valve stenosis — the MODAS Study

**DOI:** 10.64898/2026.10.26.26366513

**Authors:** Raphael Phinicarides, Kathrin Klein, Christian Jung, Bernardo S. Franklin, Ibrahim Hawwari, Victor Mauri, Jan Wrobel, Stephan Baldus, Julian A. Luetkens, Jasmin Shamekhi, Georg Nickenig, Verena Veulemans, Malte Kelm, Tobias Zeus

**Author notes:** Corresponding author: Tobias Zeus, MD, Head of Structural Heart Disease, Department of Cardiology, Pneumology and Vascular Medicine, Medical Faculty, Heinrich Heine University, University Hospital Düsseldorf, Düsseldorf, Germany.

## Abstract

**Background:** Calcific aortic stenosis (AS) is one of the most common and prognostically relevant valvular heart diseases, with a growing global burden driven by population ageing. Prediction of disease progression from moderate to severe AS, which would be instrumental for earlier diagnosis and treatment, remains challenging. At the same time, moderate AS is increasingly recognized as a heterogeneous condition associated with clinically relevant risk, particularly in patients with cardiac damage, heart failure, or rapid disease progression. Conventional echocardiographic parameters incompletely capture the biological heterogeneity underlying AS progression. The MODAS study was established to investigate biomarkers and mechanisms of AS progression and outcome using longitudinal multimodal phenotyping.

**Methods:** The MODAS Study is a prospective, multicenter, longitudinal observational cohort study enrolling adults with moderate calcific tricuspid AS at three tertiary heart valve centers in Germany. Participants undergo standardized baseline assessment including clinical phenotyping, laboratory testing, electrocardiography, transthoracic echocardiography, cardiac computed tomography and cardiac magnetic resonance imaging. Biospecimens are collected for biomarker, immunologic, proteomic, and genetic analyses, including genome-wide association studies and assessment of clonal haematopoiesis. Participants are followed annually for up to 5 years with repeat clinical assessment, laboratory testing, and echocardiography. The primary endpoint is time to progression to severe AS according to 2025 ESC/EACTS guideline criteria or aortic valve replacement for severe AS, adjudicated by the local multidisciplinary Heart Team. Secondary endpoints include all-cause mortality and additional clinical outcome trajectories. Multimodal data integration will be used to develop and validate prediction models for disease progression and outcome.

**Discussion:** The MODAS Study will provide a harmonized, deeply phenotyped longitudinal dataset to improve risk stratification in moderate AS and to identify imaging, inflammatory, and genetic markers associated with rapid progression and adverse outcomes. The study is designed to support future personalized surveillance strategies, improved selection for earlier intervention, and mechanistic understanding of calcific AS progression.

**Trial registration:** ClinicalTrials.gov NCT05851209. Registered on 9 May 2023.

## Introduction

Calcific aortic stenosis (AS) is one of the most frequent and lethal valvular heart diseases in contemporary practice, with a substantial global disease burden that is rising with ageing populations [1, 2]. Despite clear guideline frameworks for diagnosis and timing of intervention, outcomes for untreated AS remain poor, underscoring the need for earlier recognition and more precise risk stratification [3, 4]. The 2025 ESC/EACTS valvular heart disease guidelines emphasize echocardiography as the cornerstone for AS grading and recommend intervention primarily for severe AS, whereas patients with non-severe AS are generally managed with surveillance [4]. However, a major unresolved clinical challenge is that risk within non-severe AS is highly heterogeneous, and it remains unclear which patients are at increased risk of rapid progression, early myocardial damage, or adverse outcomes and may therefore benefit from closer monitoring or earlier treatment strategies.

A key unmet need is the management of moderate AS, which is increasingly recognized as a heterogeneous condition with clinically relevant risk, particularly when accompanied by LV dysfunction, heart failure (HF), or broader cardiac damage [5]. In a systematic review and meta-analysis, moderate AS was associated with higher mortality compared with no/mild AS, with risk amplification in subgroups such as symptomatic patients and those with impaired LVEF, supporting the concept that “moderate” haemodynamic grading does not equate to “moderate” biological risk [5]. Consistent with this, contemporary outcome data highlight a substantial mortality burden among patients with AS who do not undergo valve replacement, pointing to both clinical and health-system gaps in timely identification and treatment [3].

Progression of AS is highly variable across individuals, and conventional echo metrics (e.g., mean gradient, peak velocity, AVA) incompletely capture the underlying biology that drives valve calcification, myocardial remodeling, and clinical decompensation [4, 6]. Recent work has focused on estimating individual progression rates and identifying predictors of rapid progression, reinforcing the need for longitudinal cohorts with repeated standardized assessments to enable personalized risk prediction rather than population-average surveillance intervals [6, 7].

Computed tomography (CT) aortic valve calcium scoring (AVCS) has become a key adjunct when echocardiographic grading is discordant, particularly in low-flow or low-gradient states, and guideline-endorsed, sex-specific AVCS thresholds enable standardized, flow-independent adjudication of AS severity with established prognostic value [4, 8, 9]. Beyond valve anatomy, myocardial injury and fibrosis are increasingly recognized as relevant components of disease severity, and cardiac magnetic resonance (CMR) provides complementary information on myocardial remodeling for improved phenotyping and risk stratification [10, 11].

Aortic stenosis is increasingly understood as an active fibro-calcific and inflammatory disease rather than a purely degenerative process, with inflammation and myocardial remodeling contributing importantly to disease initiation and progression [12]. This evolving concept supports the investigation of immunological parameters in humans as potential markers and mediators of disease activity. Among these, platelet-driven immune responses may be of particular relevance, as experimental data indicate that platelets can modulate monocyte activation and cytokine production through direct cellular interactions and soluble mediators [13]. In addition, genetic and epigenetic mechanisms may contribute to interindividual differences in inflammatory activity and disease trajectory. In this context, clonal haematopoiesis of indeterminate potential (CHIP) has emerged as a clinically relevant modifier of cardiovascular inflammation and has also been associated with AS progression, supporting the integration of genetic and immunological profiling in the present study [4, 12].

Within this context, the MODAS study (Mechanisms Of Disease progression in Aortic Stenosis) project was established to investigate biomarkers and mechanisms of AS progression and outcome in a longitudinal human cohort, integrating morphological, functional, genetic, and immunological parameters and explicitly aiming to develop predictive models using advanced analytics including deep learning. MODAS constitutes the clinical cohort of subproject C06 of the Collaborative Research Center/Transregio TRR259 “Aortic Diseases” (Bonn–Cologne– Düsseldorf), funded by the German Research Foundation (DFG). The study is embedded within the GUARD (genes underlying aortic valve disease) network, a collaborative research platform of the participating centers that pools clinical, imaging, biobank, and genetic data from patients with bicuspid and tricuspid calcific AS to investigate the genetic and molecular determinants of disease; MODAS contributes its longitudinal moderate-AS cohort and biobank to this platform and uses its harmonized sequencing and genotyping pipelines [12, 14]. Here, we describe the rationale and design of this prospective longitudinal study (ClinicalTrials.gov: NCT05851209); the detailed study procedures, phenotyping strategy, and endpoints are provided in the Materials and Methods section to support transparent reporting and facilitate future harmonization and external validation.

## Methods

### Study design and setting

The MODAS Study is a prospective, multicenter, longitudinal observational cohort study enrolling adults with moderate, calcific (tricuspid) aortic stenosis (AS) and following them for up to 5 years. Participants are recruited at three tertiary heart valve centers in Germany (University Hospital Düsseldorf (coordinating center), University Hospital Cologne, and University Hospital Bonn) and undergo a standardized baseline phenotyping program plus annual follow-up visits for 5 years (Table 1). All three sites are Heart Team-based tertiary valve centers of the TRR259 consortium with dedicated echocardiography, cardiac CT, and CMR units.

**Table 1.** Schedule of study assessments.

| Assessment | Baseline (Visit 0) | Annual follow-up (Visits 1–5) | Clinically indicated only |
| --- | --- | --- | --- |
| Written informed consent (incl. optional genetic consent) | X |  |  |
| Structured history, medication, comorbidities, NYHA/CCS class | X | X |  |
| Events ascertainment (hospitalisation, intervention, vital status) |  | X |  |
| Clinical Frailty Scale, surgical risk scores | X | X |  |
| Physical examination and vital signs | X | X |  |
| 12-lead ECG | X | X |  |
| Standardized laboratory panel | X | X |  |
| Six-minute walk test | X | X |  |
| Transthoracic echocardiography (standardized protocol, Table 2) | X | X |  |
| Cardiac CT (non-contrast calcium scoring + coronary CTA) | X |  | X (e.g., pre-procedural CT before AVR; exploratory analyses only) |
| Cardiac MRI (cine, tissue characterization) | X |  | X (exploratory analyses only) |
| Biobanking: EDTA, citrate, heparin plasma; PBMC | X | X |  |
| Biobanking: PAXgene RNA tube, CPT tube (CHIP), germline DNA (GWAS) | X |  |  |
| Primary endpoint adjudication (Heart Team review of the predefined minimum dataset) |  | when progression to severe AS is suspected | X |

AS, aortic stenosis; AVR, aortic valve replacement; CHIP, clonal haematopoiesis of indeterminate potential; CPT, cell preparation tube; CTA, computed tomography angiography; GWAS, genome-wide association study; PBMC, peripheral blood mononuclear cells.

### Study population Eligibility

Eligible participants are adults (≥18 years) with moderate calcific AS of a tricuspid aortic valve confirmed by transthoracic echocardiography (TTE) who provide written informed consent. In accordance with the 2025 ESC/EACTS guidelines [4], moderate AS is defined by a peak aortic jet velocity (Vmax) of 3.0–4.0 m/s and/or a mean transvalvular gradient of 20–40 mmHg, with an aortic valve area (AVA) of 1.0–1.5 cmZ by the continuity equation. A haemodynamic criterion (Vmax or mean gradient within the moderate range) is mandatory for inclusion; AVA serves as a confirmatory parameter. Participants who fulfil the haemodynamic criterion but have an AVA >1.5 cmZ (e.g., because of large body size or high transvalvular flow) are enrolled, with AVA additionally assessed indexed to body surface area and in relation to flow, and are prospectively flagged in the database as “discordant AVA”. Calcific AS is defined by echocardiographic evidence of leaflet calcification with restricted cusp motion; the calcification burden is quantified at baseline by CT calcium scoring.

Patients with reduced left ventricular ejection fraction (LVEF) and patients with low-flow states (stroke volume index ≤35 mL/mZ) are eligible, since these subgroups carry the highest risk and are of particular interest for progression research. Flow status, the dimensionless velocity index (DVI), and the CT aortic valve calcium score are recorded in all participants to characterize discordant flow–gradient patterns at baseline. Discordant grading is handled as follows: (i) patients with AVA <1.0 cmZ and mean gradient <40 mmHg (possible low-flow, low-gradient severe AS) are not eligible, as they require the guideline-recommended diagnostic work-up for severe AS; (ii) patients with AVA 1.0–1.5 cmZ but Vmax <3.0 m/s and mean gradient <20 mmHg in the presence of normal flow are classified as mild AS and are not eligible; (iii) in low-flow states with gradients at the lower moderate boundary, severity is classified using the integrated approach recommended by the guidelines, combining AVA, stroke volume index, DVI, and sex-specific CT calcium score thresholds [4, 8, 15]. Discordant categories — AVA >1.5 cmZ with a moderate-range gradient, and low-flow states with gradients at the lower moderate boundary — are prespecified analysis strata, and the primary analysis is repeated in the concordant subgroup as a sensitivity analysis.

Key exclusion criteria are bicuspid aortic valve, severe AS at baseline, non-calcific AS etiologies (e.g., rheumatic or radiation-induced disease), prior aortic valve intervention, and inability to provide informed consent. Standard contraindications to the study imaging modalities as per local clinical routine apply, namely severe renal impairment or documented allergy to iodinated contrast for contrast-enhanced CT, and non-MR-conditional implants, severe claustrophobia, or contraindications to gadolinium-based contrast agents for CMR. Patients with a contraindication to a single modality are enrolled and phenotyped with the remaining modalities; the reason for the missing examination is documented.

Written informed consent is obtained before any study-specific procedure, i.e., prior to baseline blood sampling and study imaging. Consent covers clinical follow-up and the storage and use of biospecimens for biomarker analyses; consent for genetic analyses is obtained as a separate, optional item and can be declined or withdrawn at any time without affecting participation in the clinical study. Upon withdrawal, stored samples designated for genetic analysis are destroyed.

### Study procedures and follow-up schedule

Participants undergo a comprehensive baseline assessment and annual follow-up visits (Visits 1– 5, within a window of ±3 months around the scheduled date). The study assessments are performed in addition to standard-of-care surveillance for moderate AS, which comprises clinical review and TTE every 1–2 years according to current guidelines [4]. The study TTE is performed annually with a standardized measurement protocol (Table 2) that exceeds routine clinical measurement sets, and the study adds ECG, laboratory and biobank sampling, functional testing, and, at baseline, CT and CMR.

**Table 2.** Standardized transthoracic echocardiography protocol (baseline and follow-up).

| Domain | Parameters |
| --- | --- |
| Left ventricular structure and function | LVEDD, LVESD, interventricular septal thickness (mm); LVEF (%; biplane Simpson) and category (preserved / mildly / moderately / severely reduced); left atrial volume index (mL/m<sup>2</sup>) |
| Aortic stenosis and LVOT haemodynamics | Vmax (m/s); peak and mean gradient (mmHg); aortic valve VTI (cm); LVOT diameter (mm) and VTI (cm); AVA by continuity equation (cm<sup>2</sup>); stroke volume (mL) and stroke volume index (mL/m<sup>2</sup>); dimensionless velocity index |
| Other valves | Aortic regurgitation grade; mitral regurgitation grade; mean mitral gradient (mmHg); tricuspid regurgitation grade |
| Right heart | Basal RV diameter (mm); TAPSE (mm); fractional area change (%); peak TR gradient (mmHg); inferior vena cava diameter and collapsibility; estimated systolic pulmonary artery pressure (mmHg) |
| Diastolic function | E and A wave velocities (cm/s); E/A ratio; septal and lateral e' (cm/s); E/e' |
| Administrative | Examination date; free-text comment on missing or non-evaluable parameters |

AVA, aortic valve area; LVEDD/LVESD, left ventricular end-diastolic/end-systolic diameter; LVEF, left ventricular ejection fraction; LVOT, left ventricular outflow tract; RV, right ventricle; TAPSE, tricuspid annular plane systolic excursion; TR, tricuspid regurgitation; VTI, velocity–time integral.

The baseline visit (Visit 0) comprises standardized clinical, imaging, laboratory, immunological, and genetic phenotyping (Table 1):

- written informed consent;
- structured clinical history and phenotyping: demographics, cardiovascular risk factors, comorbidities, current medication, symptom status (NYHA and CCS class), Clinical Frailty Scale, and surgical risk scores;
- physical examination and vital signs;
- 12-lead electrocardiography;
- standardized laboratory panel (haematology, renal and hepatic function, lipid profile, glycaemic and thyroid parameters, cardiac and inflammatory biomarkers, coagulation, and urine analysis);
- six-minute walk test;
- transthoracic echocardiography;
- cardiac CT and cardiac magnetic resonance imaging (CMR);
- biospecimen collection for biobanking, including dedicated samples for immunological profiling, genome-wide genotyping, and clonal haematopoiesis assessment. Annual follow-up visits (Visits 1–5) include (Table 1):
- interval history and events ascertainment (symptoms, NYHA class, hospitalisations, interventions, and vital status);
- repeat clinical phenotyping including Clinical Frailty Scale;
- 12-lead electrocardiography and the standardized laboratory panel;
- biospecimen collection (plasma and peripheral blood mononuclear cells; no repeat genetic sampling);
- six-minute walk test;
- transthoracic echocardiography with the standardized protocol to track haemodynamic progression and ventricular remodeling.

Cardiac CT and CMR are not part of the scheduled follow-up assessments and are repeated only when clinically indicated (see below). Serial CT or CMR imaging is not a study objective; any analysis of clinically indicated repeat imaging is exploratory and hypothesis-generating.

### Imaging

#### Transthoracic echocardiography

TTE is performed at baseline and yearly, applying contemporary guideline definitions and recommended methods for quantifying AS severity and ventricular response [4, 16].

Examinations are performed by experienced physicians and sonographers in the echocardiography units of the participating centers on commercially available ultrasound systems. The standardized measurement protocol (Table 2) comprises: left ventricular dimensions (LVEDD, LVESD, interventricular septal thickness), LVEF (biplane Simpson) with categorization as preserved, mildly, moderately, or severely reduced, and left atrial volume index; AS haemodynamics including Vmax, peak and mean transvalvular gradient, aortic valve velocity–time integral (VTI), left ventricular outflow tract (LVOT) diameter and VTI, AVA by the continuity equation, stroke volume and stroke volume index, and DVI; grading of aortic, mitral, and tricuspid regurgitation and mean mitral gradient; right ventricular parameters (basal RV diameter, TAPSE, fractional area change, peak tricuspid regurgitation gradient, inferior vena cava assessment, and estimated systolic pulmonary artery pressure); and diastolic function (E, A, E/A ratio, septal and lateral e′, and E/e′). Global longitudinal strain and three-dimensional planimetry of the AVA are not part of the mandatory core protocol. Measurements are entered into the electronic case report form by the reading physician. Inter-observer variability will be assessed by blinded re-reading of a random subset (10%) of baseline examinations at the coordinating center, and intraclass correlation coefficients for the key AS parameters will be reported.

#### Cardiac CT

Cardiac CT is performed at baseline to enable detailed phenotyping of valve pathology (Figure 1) [8, 17]. The same acquisition protocol is used at all participating sites on ECG-gated dual-source or ≥64-slice multidetector systems (SOMATOM Definition Flash, Siemens Healthineers, Forchheim, Germany, at the coordinating center), following the acquisition standards of the

**Fig. 1.**
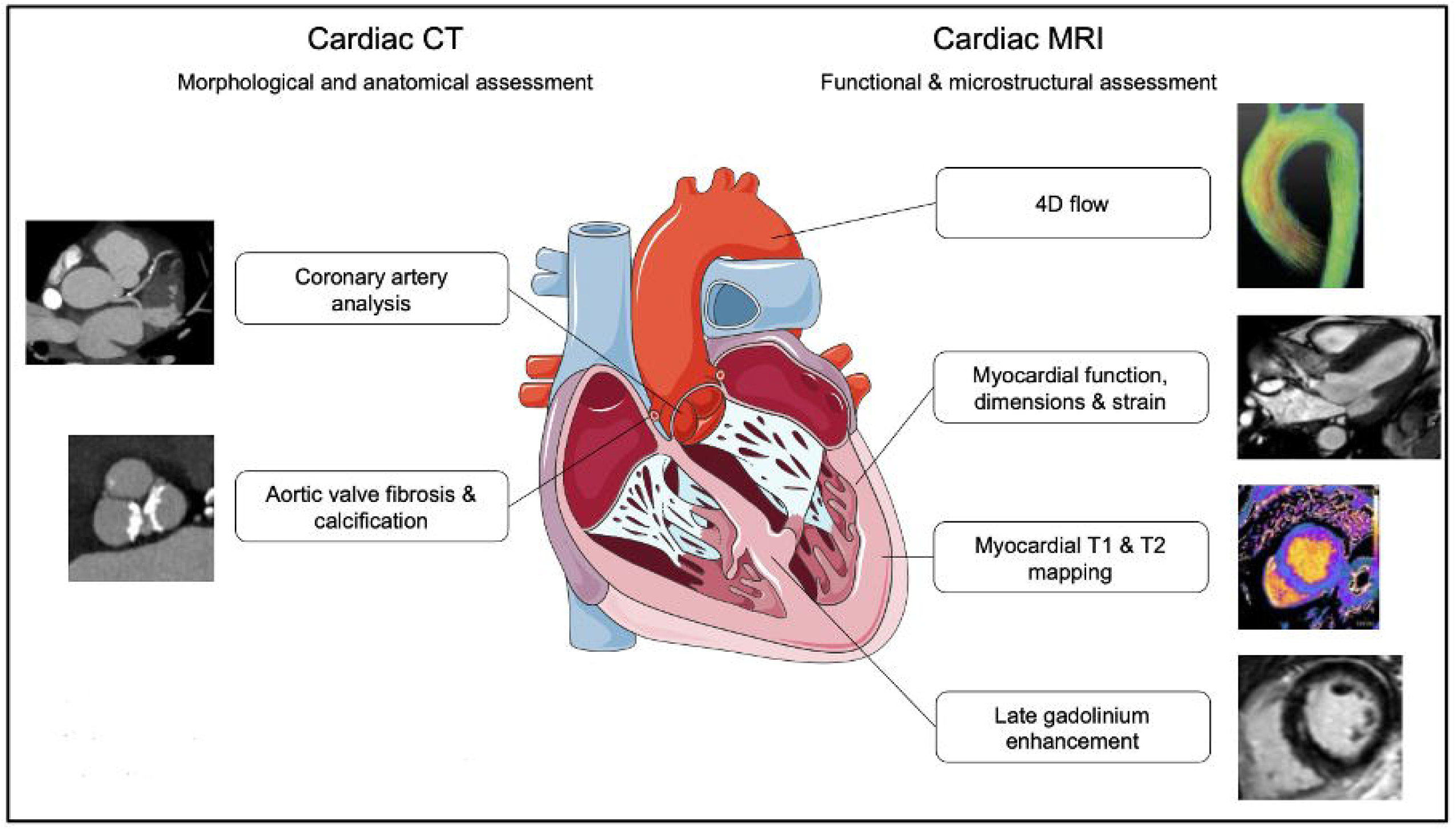
Cardiac CT and MRI biomarkers for the assessment of disease severity and outcome in patients with aortic stenosis. Alt-text: Schematic illustration of a heart in cross-section at the center, with imaging biomarkers arranged on both sides. On the left, under the heading “Cardiac CT – morphological and anatomical assessment”, two CT images with labels “Coronary artery analysis” and “Aortic valve fibrosis and calcification”. On the right, under the heading “Cardiac MRI – functional and microstructural assessment”, four MRI images with labels “4D flow”, “Myocardial function, dimensions and strain”, “Myocardial T1 and T2 mapping”, and “Late gadolinium enhancement”, each connected by a line to the corresponding cardiac structure.

Society of Cardiovascular Computed Tomography [18]. After a topogram, a non-contrast, prospectively ECG-triggered acquisition (120 kV, 3 mm slice thickness) is obtained for quantification of aortic valve and coronary calcification by the Agatston method; aortic valve calcium burden is graded against sex-specific thresholds [4, 15]. This is followed by a prospectively ECG-triggered, diastolic coronary CT angiography after intravenous injection of 100 mL of iodinated contrast (350 mg iodine/mL), covering the aortic valve, aortic root, and coronary arteries for anatomical characterization. Image quality is documented for every examination, and quantification is performed on dedicated post-processing software. Calcium quantification is performed with identical acquisition and reconstruction settings at all sites to ensure cross-site comparability.

The baseline CT constitutes the single study-related radiation exposure per participant approved by the German Federal Office for Radiation Protection (see Statement of Ethics). CT is therefore not repeated for study purposes. CT examinations performed during follow-up for clinical indications — in particular pre-procedural CT before TAVI or surgical AVR, which is standard of care and includes calcium quantification, and CT calcium scoring for discordant echocardiographic grading — are captured in the study database together with the documented indication, enabling paired analyses of calcification progression in the subset of participants with serial CT, which are prespecified as exploratory and hypothesis-generating analyses.

#### Cardiac magnetic resonance imaging

CMR is performed at baseline for standardized assessment of ventricular volumes, mass, systolic function, aortic flow, and myocardial tissue characterization (Figures 1 and 2), following the standardized protocols of the Society for Cardiovascular Magnetic Resonance [19]. The same dedicated study protocol is used at all participating sites; at the coordinating center, imaging is performed on a 1.5 T system (Achieva, Philips Healthcare, Best, The Netherlands). After survey and anatomical reference scans, ECG-triggered breath-hold cine steady-state free precession imaging is acquired in the two-, three-, and four-chamber views, the left ventricular outflow tract and aortic valve planes, and as a short-axis stack with full ventricular coverage from base to apex (repetition time/echo time shortest, flip angle 60°, spatial resolution 8 × 1.2 × 1.2 mm3, 30 phases). Four-dimensional flow imaging of the thoracic aorta is acquired for quantification of transvalvular and aortic flow. Myocardial tissue characterization comprises native T1 mapping and T2 mapping (multi-echo gradient-spin-echo) in short-axis orientation before contrast, and, after intravenous administration of gadoteric acid (Clariscan, GE Healthcare, Chicago, IL, USA; 0.2 mmol/kg body weight), three-dimensional late gadolinium enhancement imaging in long-and short-axis orientations with phase-sensitive inversion recovery reconstruction and post-contrast T1 mapping for calculation of the extracellular volume fraction using the same-day haematocrit. A three-dimensional whole-heart mDIXON acquisition is obtained for anatomical characterization of the aortic root and coronary origins. All sequences are part of the standard study protocol at all sites; examinations in which individual sequences could not be acquired (e.g., because of arrhythmia or limited breath-hold capacity) are documented, and the missing data are handled as described in the statistical analysis section. CMR is not repeated for study purposes; clinically indicated repeat CMR during follow-up is captured with its documented indication and analysed on an exploratory basis only.

**Fig. 2.**
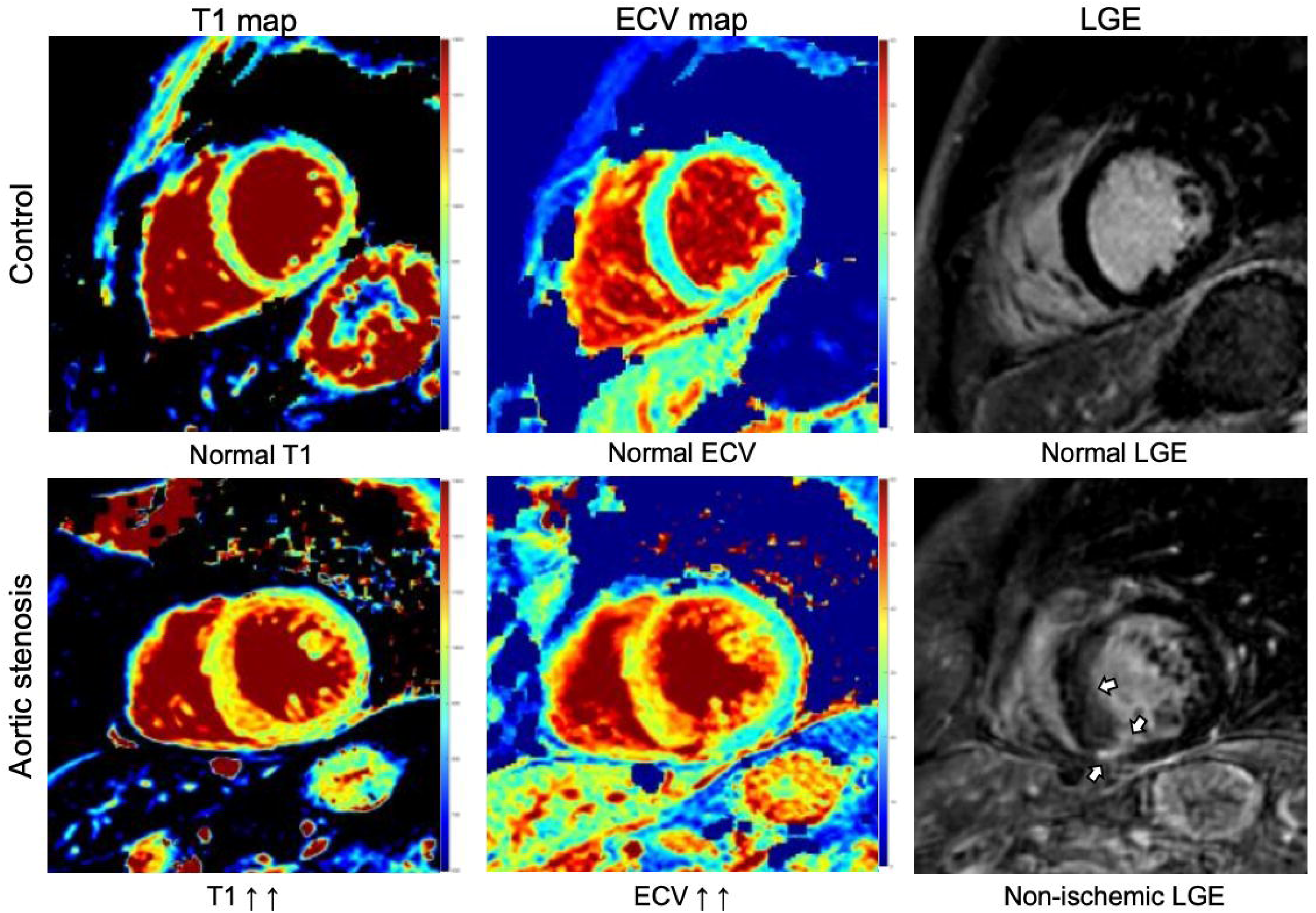
Example of multiparametric cardiac MRI parameters in a healthy control (30-year-old man) and patient (62-year-old man) with AS. The AS patient has increased myocardial T1 relaxation times and extracellular volume (ECV) values. On late gadolinium enhancement (LGE) images focal areas of replacement fibrosis are visible (arrows). Alt-text: Two rows of three short-axis cardiac MRI images. The top row, labelled “Control”, shows a colour-coded T1 map and ECV map of the left ventricle with homogeneous, predominantly blue-green (normal) values, and a greyscale late gadolinium enhancement image without enhancement, labelled “Normal T1”, “Normal ECV” and “Normal LGE”. The bottom row, labelled “Aortic stenosis”, shows the same three images from a patient: the T1 and ECV maps display a predominantly red-yellow myocardium indicating markedly elevated values (labelled “T1 ↑↑” and “ECV ↑↑”), and the late gadolinium enhancement image shows focal bright areas of non-ischemic replacement fibrosis marked by white arrows (labelled “Non-ischemic LGE”).

#### Biospecimen collection and biobanking

Peripheral venous blood is collected at baseline and follow-up using predefined collection tubes and standardized processing workflows harmonized across sites. Biospecimens are used for routine laboratory and biomarker analyses, including haematologic, biochemical, coagulation, inflammatory, lipid, endocrine, immunologic, and urine-related phenotyping, as well as for dedicated molecular analyses such as genetic profiling, genome-wide association studies (GWAS), and clonal haematopoiesis (CHIP) assessment. In addition, selected samples are collected for downstream transcriptomic and proteomic analyses where applicable. Samples are processed, aliquoted, and stored under standardized biobank conditions for subsequent analyses.

At baseline, blood is drawn into EDTA, citrate, and lithium-heparin tubes for plasma, into a PAXgene RNA tube for transcriptomic analyses, and into a cell preparation tube (CPT) for mononuclear cell isolation. Plasma is separated by centrifugation (EDTA: 3000 g for 10 min; citrate and heparin: 500 g for 10 min) and stored in 500 µL aliquots; additional EDTA whole-blood aliquots are frozen unprocessed as back-up and as a source of germline DNA. Peripheral blood mononuclear cells (PBMCs) are isolated from pooled EDTA blood by Ficoll density-gradient centrifugation (700 g for 20 min without brake), washed twice, and cryopreserved in 90% fetal calf serum/10% DMSO; mononuclear cells from the CPT tube are pelleted after centrifugation (1700 g for 20 min) and cryopreserved in the same medium. PAXgene tubes are frozen unprocessed. All aliquots are stored at −80 °C in the local biobank; cryopreserved cells are transferred to −150 °C for long-term storage. At follow-up visits, plasma and PBMC samples are collected; genetic sampling is not repeated.

Immunological profiling is performed centrally at the Institute of Innate Immunity, University of Bonn, on cryopreserved samples processed with the harmonised biobanking standard operating procedure at all sites, so that no cross-site assay variability arises. Following the methods established by the group [13], it comprises multiparameter flow-cytometric immunophenotyping of PBMCs with a focus on monocyte subsets and platelet–monocyte aggregates, ex vivo assessment of monocyte cytokine responses with and without platelets, and plasma cytokine profiling. Standardised gating strategies and inter-run reference controls are applied, and the analytical batch is recorded and included as a covariate in all analyses. The primary immunological readouts for the MODAS analyses will be prespecified in the statistical analysis plan before database lock, and immunological variables enter only the exploratory tier of the prediction models (see Statistical analysis).

CHIP is assessed by targeted next-generation sequencing of DNA from peripheral blood using a panel of 48 recurrently mutated clonal haematopoiesis driver genes (Illumina NovaSeq 6000; mean sequencing depth approximately 20,000×), with CHIP defined as the presence of a driver mutation at a variant allele frequency ≥2% (≥4% for X-linked genes in men), using the pipeline established within the GUARD cohort [12]. In the analyses, CHIP is modelled primarily as a binary variable (presence of any driver mutation at VAF ≥2%); clone size (largest VAF, continuous and ≥10%) and gene-specific analyses (DNMT3A, TET2, other genes) are secondary and exploratory. Genome-wide genotyping is performed on germline DNA within the GUARD consortium using a genome-wide array with subsequent imputation. MODAS is not powered for stand-alone discovery GWAS; genetic data will be contributed to consortium-level meta-analyses and used for polygenic risk score analyses in the MODAS cohort. Multiple-testing correction is described in the statistical analysis section.

### Study endpoints

#### Primary endpoint

The primary endpoint is the time from enrolment to the first occurrence of either (a) progression from moderate to severe AS or (b) aortic valve replacement (AVR; TAVI or surgical AVR) performed for severe AS. Severe AS is defined according to the 2025 ESC/EACTS guidelines [4]: Vmax ≥4.0 m/s or mean gradient ≥40 mmHg; or AVA <1.0 cmZ (indexed AVA <0.6 cmZ/mZ) in low-flow, low-gradient states when severity is confirmed by the guideline-recommended integrated assessment, including sex-specific CT calcium score thresholds and, in patients with reduced LVEF, low-dose dobutamine stress echocardiography where applicable.

Endpoint adjudication follows a predefined process. Whenever progression to severe AS is suspected — at a scheduled study TTE or on clinical grounds between visits — the treating study investigator presents the patient with all available imaging findings to the local multidisciplinary Heart Team (cardiology, cardiac surgery, and anaesthesiology). The minimum dataset for Heart Team review comprises current symptom status and NYHA class, the 12-lead ECG, the index TTE with the complete standardized AS dataset (Vmax, mean gradient, AVA, stroke volume index, DVI, and LVEF), the baseline CT calcium score and any clinically indicated repeat CT, the natriuretic peptide level, and the comorbidity and frailty assessment. The Heart Team adjudicates on the basis of the clinical record and is therefore blinded by design to all research biomarker, immunological, and genetic data, which do not enter the clinical record and are not analysed before database lock; it is not blinded to the clinical baseline imaging, which is part of routine care and required for the severity assessment. After review of the findings and the clinical history, the Heart Team confirms or rejects the diagnosis of severe AS and issues a treatment recommendation according to the ESC/EACTS guidelines [4]. The Heart Team decision, the date of the index imaging study on which it is based, and — if applicable — the indication and date of AVR are documented in the study database. A confirmed diagnosis of severe AS counts as a primary endpoint event with the date of the index imaging study as event date; AVR counts as a primary endpoint event only if it is preceded by a Heart Team-confirmed diagnosis of severe AS. AVR performed without confirmed severe AS (for example, replacement of a still-moderate valve during coronary bypass or other cardiac surgery, or aortic valve surgery for infective endocarditis) is not counted as a primary endpoint event; such participants are censored at the date of surgery and analysed separately in sensitivity analyses. The two components of the composite endpoint are additionally analysed separately as secondary endpoints. Because the decision to intervene is not standardized by the protocol, the proportion of participants with confirmed severe AS who undergo AVR, and the time from confirmation to AVR, are monitored and reported annually by site as quality indicators; at the current data cutoff this proportion was 76% at the coordinating center and 62% at each of the two other sites. Center is a stratification factor in all time-to-event analyses, and the sensitivity analysis treating AVR as a censoring event is independent of site-level intervention thresholds.

#### Secondary endpoints

Secondary endpoints include major clinical outcomes such as all-cause mortality, and additional clinical events and trajectories (including symptom progression, heart-failure–related outcomes, and procedure-related pathways) as predefined in Table 3 and captured systematically at each follow-up visit through structured events ascertainment (hospitalisations, interventions, vital status), symptom and frailty assessment, the six-minute walk test, and the standardized echocardiography protocol. Vital status and date of death are verified through hospital records and, where necessary, contact with the treating physician or the population registry. Rapid haemodynamic progression is prespecified as an annual increase in Vmax of ≥0.3 m/s, the threshold derived from the natural-history study of Rosenhek et al. and adopted by the ESC/EACTS guidelines to define rapid progression [4, 20]. Serial CT and CMR parameters are listed in Table 3 as exploratory endpoints only.

**Table 3.** Primary and secondary endpoints.

| **Endpoint** | **Definition / assessment** |
| --- | --- |
| Primary endpoint | Time from enrolment to the first occurrence of Heart Team-confirmed progression to severe AS (2025 ESC/EACTS criteria) or AVR (TAVI or surgical AVR) performed for severe AS. Primary estimand: 5-year cumulative incidence in the presence of the competing risk of death (Aalen–Johansen); confirmatory association analyses: cause-specific hazards (Cox, stratified by center) |
| Components of the primary endpoint (secondary) | Progression to severe AS by imaging criteria, with AVR treated as censoring event; AVR for severe AS |
| All-cause mortality | Death from any cause and date of death; vital status verified at each visit and through hospital records, treating physicians, or the population registry |
| Any aortic valve replacement | AVR irrespective of indication, with date and type of procedure (TAVI or surgical AVR); AVR without Heart Team-confirmed severe AS (e.g., during coronary bypass surgery or for infective endocarditis) analysed in sensitivity analyses |
| Symptom progression | Increase in NYHA class by $\geq 1$ or new angina (CCS class) or syncope attributed to AS at a scheduled visit |
| Heart failure-related outcomes | Hospitalisation for heart failure and new or worsening heart failure symptoms, ascertained from the structured interval history and hospital records |
| Other cardiovascular hospitalisations and interventions | Unplanned cardiovascular hospitalisations and cardiac interventions during follow-up, ascertained from the structured interval history |
| Functional and frailty trajectory | Change in six-minute walk distance and in the Clinical Frailty Scale from baseline |
| Haemodynamic progression | Annualized change in Vmax, mean gradient, and AVA on the standardized study TTE; rapid progression prespecified as an increase in Vmax $\geq 0.3$ m/s per year, threshold derived from Rosenhek et al. [20] |
| Cardiac remodeling (echocardiography) | Change in LVEF, LV dimensions, left atrial volume index, diastolic function, and right ventricular function on serial study TTE |
| Serial CT and CMR parameters (exploratory) | Change in CT aortic valve calcium score and in CMR tissue characterization (native T1, ECV, LGE) in participants with clinically indicated repeat imaging; not a study objective, exploratory and hypothesis-generating only |

AS, aortic stenosis; AVA, aortic valve area; AVR, aortic valve replacement; CCS, Canadian Cardiovascular Society; ECV, extracellular volume fraction; LGE, late gadolinium enhancement; LV, left ventricular; LVEF, left ventricular ejection fraction; TTE, transthoracic echocardiography.

#### Sample size calculation

The planned sample size is 938 participants. This estimate was based on the expected precision of the incidence of progression to haemodynamically significant severe aortic stenosis according to current ESC guideline criteria over the 5-year follow-up period. In longitudinal cohorts of moderate AS with serial echocardiography, progression to severe AS has been reported in 39% of patients over a mean follow-up of 4.8 years [7] and in 47% of patients with mild or moderate AS, with 56% progressing within 5 years [21]; our own pilot data showed progression of calcific aortic valve disease in 37% of patients with aortic valve sclerosis within 4 years [22]. Because these rates were derived from populations that differ from the MODAS cohort in baseline severity, a conservative 5-year event rate of 40% was assumed. MODAS is a descriptive and model-development cohort without a comparator group; the sample size was therefore based on the precision of the primary estimate rather than on power for a hypothesis test. The required number of participants was derived from the precision of the Kaplan–Meier estimate of the 5-year cumulative incidence of the primary endpoint (Greenwood variance), allowing for 20% censoring before 5 years through death — approximately 18% at 5 years in comparable moderate-AS populations [7] — or loss to follow-up. With p = 0.40 and a two-sided α of 0.05, 750 participants with complete follow-up yield a half-width of the 95% confidence interval of 3.5 percentage points under the binomial approximation, n = zZ·p·(1 − p)/dZ (z = 1.96, d = 0.035, n ≈ 752), and 938 enrolled participants yield a half-width of approximately 3.3 percentage points for the Kaplan–Meier estimate under the censoring assumption; the planned enrolment is therefore 750/0.8 = 938 participants. In a sensitivity analysis across plausible 5-year event rates of 30%, 40%, and 50%, the expected half-width of the 95% confidence interval of the Kaplan– Meier estimate is 3.1, 3.3, and 3.4 percentage points, with approximately 255, 340, and 425 expected events, respectively, so that the planned sample size provides adequate precision over the entire plausible range. Based on the 750 completers, approximately 300 participants with progression to severe AS and 450 participants with stable moderate AS are expected to be available for analysis. For the analysis of predictors, approximately 300 events provide 80% power (two-sided α of 0.05) to detect a cause-specific hazard ratio of 1.38 for a binary predictor with 50% prevalence and of 1.45 for a predictor with 25% prevalence.

For the development of multivariable prediction models, the sample size is driven by the number of events relative to the number of candidate predictor parameters. With approximately 300 events, a conservative ratio of at least 10 events per candidate parameter supports stable estimation of models with up to about 30 candidate predictor parameters in the prespecified (Tier 1) clinical–imaging model, which is sufficient for a clinically oriented model integrating demographic, echocardiographic, CT, CMR, and selected biomarker variables. Formal sample-size criteria for prediction models (minimizing overfitting, achieving a small absolute difference between apparent and optimism-adjusted performance, and precise estimation of the overall risk) will be verified according to Riley et al. [23] before model development, and high-dimensional immunological and genetic data will be entered only through dimension-reducing or penalized approaches as described below.

#### Statistical analysis

Continuous variables will be summarized using appropriate measures of central tendency and dispersion, and categorical variables as counts and percentages. The primary analysis population (full analysis set) comprises all enrolled participants who fulfil the eligibility criteria and have a baseline TTE; given the observational design, no intention-to-treat or per-protocol distinction applies.

The primary endpoint will be analysed as a time-to-event outcome. The primary estimand is the 5-year cumulative incidence of the primary endpoint in the presence of the competing risk of death, estimated with the Aalen–Johansen estimator; Kaplan–Meier estimates are reported for descriptive comparison only. For the association of baseline characteristics with progression, the primary (confirmatory) analysis uses cause-specific hazard ratios from Cox proportional-hazards models stratified by study center, because the aetiological question concerns the rate of progression among participants at risk. Fine–Gray subdistribution hazard models are supportive analyses and are used where the estimand is absolute risk, namely for the prediction models.

Prespecified sensitivity analyses comprise (i) progression to severe AS by imaging criteria only, with AVR treated as a censoring event; (ii) a composite including all AVR irrespective of indication; (iii) exclusion of participants with AVR for non-AS indications; and (iv) restriction to participants with concordant grading at baseline. To avoid immortal time bias, exposures that arise during follow-up will be modelled as time-dependent covariates or using landmark analyses. Longitudinal echocardiographic and biomarker trajectories will be analysed with linear mixed-effects models with random intercepts and slopes for participants and a random effect for center; center will otherwise be handled by stratification or as a random effect in all multivariable models.

Missing data will be addressed according to a prespecified strategy. Missingness will be quantified and its pattern described for every variable. For baseline predictors, multiple imputation by chained equations (20 imputed datasets, including the outcome and auxiliary variables in the imputation model) will be used under the missing-at-random assumption, with complete-case analyses as sensitivity analyses. In the primary time-to-event analysis, outcome status and event dates are not imputed; for the prediction models the outcome is likewise taken as observed, and imputation applies to predictors only. Missing serial imaging data will not be imputed; analyses of imaging progression will be restricted to participants with paired examinations, with sensitivity analyses comparing baseline characteristics of participants with and without repeat imaging to assess selection. Differential follow-up intensity will be addressed by site-level monitoring of visit adherence and by sensitivity analyses restricted to participants with complete scheduled follow-up.

Prediction models for progression will be developed in a prespecified two-tier hierarchy. Tier 1 (prespecified, confirmatory) is a parsimonious clinical–imaging model with at most 30 candidate predictor parameters, prespecified on the basis of clinical knowledge and the literature (demographic, clinical, laboratory, echocardiographic, CT, and CMR variables, and ECG-derived features including, where available, artificial-intelligence-based ECG markers of AS [24]); univariable screening will not be used for variable selection. The Tier 1 model will be fitted with penalized Cox regression (LASSO or elastic net) and validated internally by bootstrap optimism correction; its discrimination (C-index), calibration (calibration plots and slope), and clinical utility (decision-curve analysis) constitute the prespecified primary prediction result. Tier 2 (exploratory, hypothesis-generating) adds immunological, CHIP, and genetic variables and complementary machine-learning algorithms (e.g., random forests and gradient boosting) for multimodal risk prediction. Overfitting in Tier 2 is controlled by (i) dimension reduction or penalization of high-dimensional variable blocks before entry, (ii) nested cross-validation with center-stratified resampling, in which imputation, feature selection, and hyperparameter tuning are repeated within each outer fold, (iii) comparison of every Tier 2 model with the Tier 1 model on identical resamples, so that incremental value is tested against the clinical model, and (iv) explicit labelling as exploratory in all reports. Tier 2 models will not be presented as validated risk tools without external validation, which is planned in independent cohorts within the GUARD network. For exploratory analyses of immunological and CHIP variables, the false discovery rate will be controlled with the Benjamini–Hochberg procedure; for genome-wide analyses, the conventional genome-wide significance threshold (p < 5 × 10⁻⁸) applies. Model development and reporting will follow the TRIPOD+AI statement for all prediction models, whether based on regression or machine learning [25]. A detailed statistical analysis plan will be finalized before database lock and will be made publicly available via the trial registry (ClinicalTrials.gov NCT05851209) before the primary analysis is performed. Analyses will be performed with R and Python.

#### Data capture, quality assurance, and governance

Clinical and study data are captured in a standardized electronic data capture system (REDCap, Research Electronic Data Capture) hosted at the coordinating center, with predefined data elements and a common data dictionary, harmonized across sites, and subject to automated range and consistency checks at data entry. Regular central data monitoring is performed by the coordinating center, including completeness and plausibility checks and query management with the sites; the system maintains a full audit trail. Data quality across sites is further assessed by central statistical monitoring of key variables (completeness, distributions, and outliers by site), including the proportion of participants with confirmed severe AS who undergo AVR and the time from confirmation to AVR by site.

#### Ethics and consent

The study is conducted in accordance with the Declaration of Helsinki and applicable data protection regulations. The protocol was approved by the ethics committees of all participating sites (lead ethics committee: Medical Faculty of Heinrich Heine University Düsseldorf, study number 2022-2236; Bonn, reference 18/19; Cologne, reference 22-1458_2), and the study-related radiation exposure was approved by the German Federal Office for Radiation Protection (approval number ZD 3-22464/2023-022-G); full details are given in the Statement of Ethics. All participants provide written informed consent covering clinical follow-up and storage/use of biospecimens for biomarker and genetic analyses, with a separate optional consent for genetic analyses as described above.

#### Reporting guideline

This study protocol is reported in accordance with the STROBE (Strengthening the Reporting of Observational Studies in Epidemiology) statement for cohort studies. The completed STROBE checklist is provided as online supplementary material.

#### Study status

This manuscript describes the current study protocol for the MODAS study. Recruitment began in July 2023. All three sites are actively recruiting. As of the data cutoff of 28 September 2026, 554 participants had been enrolled (Düsseldorf n = 205, Bonn n = 185, Cologne n = 164), corresponding to an average enrolment of approximately 190 participants per year since 2024; the mean age of enrolled participants is 76 years (range 46–96), and 36% are women. The first participants have returned for 1-, 2-, and 3-year follow-up visits; at the coordinating center, 183 participants have completed the 1-year follow-up visit to date. As of the same data cutoff, 46 participants had reached the primary endpoint of Heart Team-confirmed progression to severe AS, of whom 31 had undergone AVR (26 TAVI, 4 surgical AVR, 1 procedure type pending documentation), and 22 participants had died. At the current enrolment rate, the target sample size of 938 is expected to be reached well before the planned completion of recruitment in July 2031. These figures are reported to document recruitment progress and feasibility only; they do not constitute study results, and no outcome analyses have been performed.

## Discussion

Moderate aortic stenosis (AS) is not a benign ‘watch-and-wait’ condition; multiple datasets show a meaningful burden of death and heart-failure events across the AS spectrum, and outcomes worsen as cardiac damage accrues [3–5]. Current guidelines still recommend intervention primarily once AS becomes severe or when other surgical indications coexist, leaving a large group of patients in whom risk is real but the timing of action remains uncertain [4]. This uncertainty is reinforced by the marked heterogeneity of disease progression and the limited ability of routine follow-up intervals to capture individual risk trajectories [6].

Against this background, the MODAS Study has been established as a large prospective multicenter cohort of patients with moderate AS, designed to provide deep longitudinal phenotyping through harmonized clinical assessment, multimodality imaging, immunological profiling, and genetic analyses. By integrating these complementary data domains, this study aims to identify mechanisms and predictors of rapid progression and adverse outcomes and to enable more individualized risk stratification in moderate AS [14].

The emphasis on standardized echocardiography plus advanced imaging is clinically motivated: CT-derived aortic valve calcium metrics provide diagnostic and prognostic information and can refine disease staging when echocardiographic parameters are discordant or when remodeling appears disproportionate [4, 8]. Furthermore, myocardial substrate matters: in the EVOLVED trial, 224 patients with asymptomatic severe AS and CMR evidence of myocardial fibrosis (late gadolinium enhancement) were randomized to early AVR or guideline-directed conservative management; early intervention did not significantly reduce the primary endpoint of all-cause death or unplanned AS-related hospitalization, but it reduced symptom burden and unplanned hospitalizations, and the trial established CMR-based fibrosis phenotyping as a feasible tool to identify a biologically higher-risk stratum [11]. This supports the inclusion of CMR tissue characterization in MODAS as a mechanistically anchored marker of myocardial vulnerability. A key differentiator of MODAS is the explicit linkage of imaging phenotypes to systems biology, reflecting growing evidence that inflammation and immune dysregulation are not just epiphenomena in calcific AS.

In particular, CHIP has emerged as a plausible driver of inflammatory amplification and has been associated with AS progression and adverse outcomes in contemporary cohorts, including the GUARD cohort of the participating centers [12, 26, 27]; embedding CHIP and immune profiling into a prospective longitudinal design creates the opportunity to move from association toward testable mechanistic pathways and actionable risk stratification.

From a translational standpoint, MODAS is aligned with the ongoing randomized trials of earlier intervention in moderate AS: the PROGRESS trial (A Prospective, Randomized, Controlled Trial to Assess the Management of Moderate Aortic Stenosis by Clinical Surveillance or Transcatheter Aortic Valve Replacement; ClinicalTrials.gov NCT04889872) and the Evolut EXPAND TAVR II Pivotal Trial (ClinicalTrials.gov NCT05149755) compare TAVI with clinical surveillance or guideline-directed management in at-risk patients with moderate AS, and the completed TAVR UNLOAD trial in 178 patients with moderate AS and heart failure with reduced ejection fraction found TAVI safe and associated with greater quality-of-life improvement, but not superior for its hierarchical primary endpoint, with substantial crossover to TAVI because of AS progression [28]. The “earlier is better” hypothesis therefore depends on better staging, enrichment, and patient selection [29] — precisely the “who progresses fast?” question that prospective phenotyping cohorts such as MODAS are designed to answer.

Methodologically, recent work shows that AS progression can be predicted with meaningful accuracy from clinical and echocardiographic data, machine-learning models can reach high discrimination, and artificial-intelligence analysis of the 12-lead ECG can detect AS and track its trajectory years before intervention [24]; yet clinical translation hinges on transparent reporting and validation [6, 21, 25]. Prediction modeling in MODAS therefore follows a prespecified two-tier plan that separates a confirmatory clinical–imaging model from exploratory multimodal and machine-learning models, with penalized regression, internal validation, planned external validation, and reporting under TRIPOD+AI [25].

Several limitations are inherent to the present study design and must be acknowledged upfront. Observational cohorts cannot establish causality, and residual confounding remains a structural concern even with careful adjustment, particularly when clinical decisions (e.g., timing of AVR/TAVI) influence endpoints [30, 31]. Although all sites are Heart Team-based tertiary centers applying the same ESC/EACTS guidelines, the decision to perform AVR is not standardized by the study protocol and may vary across sites and over time; the Heart Team-based adjudication and the prespecified sensitivity analyses treating AVR as a censoring event are intended to limit, but cannot eliminate, the resulting ascertainment bias. A related concern is immortal time bias, since participants must survive and remain under surveillance to undergo AVR; this is addressed by time-to-event analysis with time-dependent modelling of exposures arising during follow-up and by competing-risk analyses with death as a competing event.

In addition, multimodal, real-world longitudinal studies are exposed to missing data and differential follow-up intensity; recent methodological work highlights that handling missingness for prediction is non-trivial and must be prespecified to avoid biased performance estimates [32, 33]. The restriction of CT and CMR to baseline, which follows from the radiation protection approval and the resources available for an observational cohort of this size, limits the assessment of serial changes in valve calcification and myocardial fibrosis to the subset of participants with clinically indicated repeat imaging; because clinically indicated imaging is more likely in patients with rapid progression, serial imaging is not a study objective, and analyses of serial imaging are exposed to reverse causation and are prespecified as exploratory and hypothesis-generating, with the indication for each repeat examination documented and modelled. The prespecified missing-data strategy and the sensitivity analyses restricted to participants with complete scheduled follow-up address differential follow-up intensity at the analysis level.

Finally, recruitment at tertiary valve centers may enrich for comorbidity and referral bias, potentially limiting generalizability to lower-acuity community populations [30]. This spectrum bias will be assessed by comparing the baseline characteristics of the MODAS cohort with those of published population-based and registry cohorts of moderate AS, and the applicability of any prediction model to lower-risk populations will require external validation.

In summary, the MODAS Study (ClinicalTrials.gov: NCT05851209) is positioned to deliver a clinically relevant framework for individualized risk stratification in moderate AS by integrating longitudinal imaging, biomarkers, and mechanistic immune/genetic profiling. If successful, its outputs should directly inform (i) follow-up interval personalization, (ii) trial enrichment strategies for early-intervention studies, and (iii) biological target discovery for disease-modifying therapies in calcific AS [4, 25].

## Supporting information

Strobe Checklist

## Statements

## Acknowledgement

The authors thank all investigators, study coordinators, and participating centers involved in the TRR259/MODAS Study project. MODAS is the clinical cohort of subproject C06 of the Collaborative Research Center/Transregio TRR259 “Aortic Diseases” network (Bonn–Cologne– Düsseldorf) supported by the German Research Foundation (DFG).

## Statement of Ethics

The study was approved by the Ethics Committee of the Medical Faculty of Heinrich Heine University Düsseldorf, Düsseldorf, Germany, acting as the lead (first-voting) ethics committee for this multicenter study (study number 2022-2236; approval dated 14 June 2023), and by the Ethics Committee of the Medical Faculty of the Rheinische Friedrich-Wilhelms-University of Bonn, Bonn, Germany (reference number 18/19; approval dated 16 March 2023). The participating center in Cologne additionally obtained approval from the Ethics Committee of the Medical Faculty of the University of Cologne, Cologne, Germany (reference number 22-1458_2). The application of ionising radiation for study purposes was approved by the German Federal Office for Radiation Protection (Bundesamt für Strahlenschutz; approval number ZD 3-22464/2023-022-G). The study is conducted in accordance with the Declaration of Helsinki and applicable data protection regulations. Written informed consent to participate in the study, including clinical follow-up and the storage and use of biospecimens for biomarker and genetic analyses, was obtained from all participants; consent for genetic analyses is obtained as a separate optional item.

## Conflict of Interest Statement

R.P., K.K., B.S.F., I.H., J.W., J.S., and J.A.L. declare no conflicts of interest. C.J. has received consulting fees from Janssen and lecture honoraria from Bristol-Myers Squibb, Edwards Lifesciences, and Daiichi Sankyo. V.M. has received speaker honoraria and travel support from Edwards Lifesciences. S.B. has received research grants, lecture honoraria, and consultancy or advisory fees from Abbott, Edwards Lifesciences, JenaValve, Medtronic, and Boston Scientific, and personal fees from Bayer, Pfizer, Boehringer Ingelheim, and Daiichi Sankyo. G.N. has received research funding, lecture honoraria, and advisory board fees from Abbott, Edwards Lifesciences, Medtronic, St. Jude Medical, Abiomed, and Boston Scientific. V.V. and T.Z. have received speaker, consulting, and/or study honoraria and travel support from Edwards Lifesciences, Medtronic, and Boston Scientific. M.K. has received institutional grant support and/or personal fees from Philips, Abbott, Medtronic, Boston Scientific, Boehringer Ingelheim, Daiichi Sankyo, Amgen, Ancora Heart, and B. Braun. All relationships are outside the submitted work. No author has received funding from device or pharmaceutical manufacturers for the MODAS Study, which is funded exclusively by the German Research Foundation (DFG, TRR259).

## Funding Sources

This work was conducted within the Collaborative Research Center/Transregio TRR259 “Aortic Diseases”, funded by the German Research Foundation (Deutsche Forschungsgemeinschaft, DFG). The funding body had no role in the design of the study and will have no role in data collection, analysis, interpretation of data, or writing of the manuscript.

## Author Contributions

RP drafted the manuscript. KK, CJ, BSF, IH, VM, JW, SB, JAL, JS, GN, VV, MK, and TZ contributed to the study design and critically revised the manuscript. All authors read and approved the final manuscript.

## Data Availability Statement

The datasets generated and/or analyzed during the current study are not publicly available. Individual participant data cannot be deposited in a public repository, because the scope of the informed consent and European and German data protection law (GDPR, BDSG) do not permit public release of patient-level data from this cohort. De-identified individual participant data and the data dictionary may be made available to qualified researchers on reasonable request after completion of the study and publication of the primary analyses (anticipated 2032/2033), subject to a data-sharing agreement, approval by the study steering committee, and institutional data protection regulations; requests for collaborative analyses within the TRR259/GUARD network may be considered earlier. The statistical analysis code will be made publicly available in a public repository at the time of publication of the primary analyses.

## Declaration of Generative AI Use

During the preparation of this manuscript, the authors used a generative AI tool (Claude, Anthropic) for language editing and structuring of the text. All content was reviewed and edited by the authors, who take full responsibility for the content of the publication. AI tools were not used for data analysis.

## Notes

### Clinical Trial

NCT05851209

### Clinical Protocols

https://clinicaltrials.gov/study/NCT05851209

### Author Declarations

The Ethics Committee of the Medical Faculty of Heinrich Heine University Duesseldorf gave ethical approval for this work (lead ethics committee of the multicenter study; study number 2022-2236; approval dated 14 June 2023). The Ethics Committee of the Medical Faculty of the Rheinische Friedrich-Wilhelms-University of Bonn gave ethical approval for this work (reference number 18/19; approval dated 16 March 2023). The Ethics Committee of the Medical Faculty of the University of Cologne gave ethical approval for this work (reference number 22-1458_2). The study-related radiation exposure was approved by the German Federal Office for Radiation Protection (Bundesamt fuer Strahlenschutz; approval number ZD 3-22464/2023-022-G).

