## Supplementary material for "Study protocol for a prospective multicenter longitudinal cohort study on progression of moderate aortic valve stenosis — the MODAS Study": Strobe Checklist

**STROBE Statement — Checklist of items that should be included in reports of cohort studies**

*Manuscript: Study protocol for a prospective multicenter longitudinal cohort study on progression of moderate aortic valve stenosis — the MODAS Study. As this manuscript is a study protocol, the results items of the checklist are marked as not applicable; the corresponding sections will be reported in the future publications of study results.*

| **Section** | **Item** | **Recommendation** | **Location in manuscript** |
| --- | --- | --- | --- |
| Title and abstract | 1a | Indicate the study design with a commonly used term in the title or the abstract | Title page; Abstract (prospective multicenter longitudinal cohort study) |
|  | 1b | Provide in the abstract an informative and balanced summary of what was done and what was found | Abstract |
| Background/rationale | 2 | Explain the scientific background and rationale for the investigation being reported | Introduction |
| Objectives | 3 | State specific objectives, including any prespecified hypotheses | Introduction (final paragraph); Methods — Study endpoints |
| Study design | 4 | Present key elements of study design early in the paper | Abstract; Methods — Study design and setting |
| Setting | 5 | Describe the setting, locations, and relevant dates, including periods of recruitment, exposure, follow-up, and data collection | Methods — Study design and setting; Study status |
| Participants | 6a | Give the eligibility criteria, and the sources and methods of selection of participants; describe methods of follow-up | Methods — Study population (Eligibility); Study procedures and follow-up schedule |
|  | 6b | For matched studies, give matching criteria and number of exposed and unexposed | Not applicable — no matched design |
| Variables | 7 | Clearly define all outcomes, exposures, predictors, potential confounders, and effect modifiers; give diagnostic criteria, if applicable | Methods — Study procedures; Imaging; Biospecimen collection; Study endpoints; Table 1 |
| Data sources/measurement | 8 | For each variable of interest, give sources of data and details of methods of assessment (measurement); describe comparability of assessment methods across groups | Methods — Imaging; Biospecimen collection and biobanking; Data capture, quality assurance, and governance |
| Bias | 9 | Describe any efforts to address potential sources of bias | Methods — Data capture, quality assurance, and governance; Discussion (limitations) |
| Study size | 10 | Explain how the study size was arrived at | Methods — Sample size calculation |
| Quantitative variables | 11 | Explain how quantitative variables were handled in the analyses; describe which groupings were chosen and why | Methods — Statistical analysis |
| Statistical methods | 12a | Describe all statistical methods, including those used to control for confounding | Methods — Statistical analysis |
|  | 12b | Describe any methods used to examine subgroups and interactions | Methods — Statistical analysis |
|  | 12c | Explain how missing data were addressed | Methods — Statistical analysis; Discussion |
|  | 12d | Explain how loss to follow-up was addressed | Methods — Statistical analysis |
|  | 12e | Describe any sensitivity analyses | Methods — Statistical analysis |
| Participants (results) | 13 | Report numbers of individuals at each stage of study; consider use of a flow diagram | Not applicable — study protocol; results are not yet available |
| Descriptive data | 14 | Give characteristics of study participants and information on exposures and potential confounders | Not applicable — study protocol; results are not yet available |
| Outcome data | 15 | Report numbers of outcome events or summary measures over time | Not applicable — study protocol; results are not yet available |
| Main results | 16 | Give unadjusted estimates and, if applicable, confounder-adjusted estimates and their precision | Not applicable — study protocol; results are not yet available |
| Other analyses | 17 | Report other analyses done, e.g. analyses of subgroups and interactions, and sensitivity analyses | Not applicable — study protocol; results are not yet available |
| Key results | 18 | Summarise key results with reference to study objectives | Not applicable — study protocol; results are not yet available |
| Limitations | 19 | Discuss limitations of the study, taking into account sources of potential bias or imprecision | Discussion (limitations paragraphs) |
| Interpretation | 20 | Give a cautious overall interpretation of results considering objectives, limitations, multiplicity of analyses, and other relevant evidence | Discussion |
| Generalisability | 21 | Discuss the generalisability (external validity) of the study results | Discussion (recruitment at tertiary valve centers) |
| Funding | 22 | Give the source of funding and the role of the funders for the present study | Statements — Funding Sources |
